# The Laro Kwo Project: A train the trainer model combined with mobile health technology for Community Health Workers in Northern Uganda

**DOI:** 10.1101/2022.10.26.22281557

**Authors:** Daniel Ebbs, Oyoo Benson, Stanton Jasicki, Sarah McCollum, Michael Cappello

## Abstract

Community Health Workers (CHWs) in low and middle income countries (LMICs) provide invaluable health resources to their community members. Best practices for developing and sustaining CHW training programs in LMICs have yet to be defined using rigorous standards and measures of effectiveness. With the expansion of digital health to LMICs, few studies have evaluated the role of participatory methodologies combined with the use of mobile health (mHealth) for CHW training program development. We completed a three-year prospective observational study aligned with the development of a community-based participatory CHW training program in Northern Uganda. Twenty-five CHWs were initially trained using a community participatory training methodology combined with mHealth and a train-the-trainer model. Medical skill competency exams were evaluated after the initial training and annually thereafter to assess retention with use of mHealth. After three years, CHWs who advanced to trainer status redeveloped all program materials using a mHealth application and trained a new cohort of 25 CHWs. Implementation of this methodology coupled with longitudinal mHealth training demonstrated an improvement in medical skills over three years among the original cohort of CHWs. Further, we found that the train-the-trainer model with mHealth was highly effective, as the new cohort of 25 CHWs trained by the original CHWs exhibited higher scores when tested on medical skill competencies. The combination of mHealth and participatory methodologies can facilitate the sustainability of CHW training programs in LMIC. Further investigations should focus on comparing specific mHealth modalities for training and clinical outcomes using similar combined methodologies.

## Introduction

In remote regions of low-to-middle income countries (LMICs), access to healthcare is often limited, with significant disparity across socioeconomically disadvantaged communities. The added cost of health services for families living in extreme poverty is especially challenging when issues like food insecurity dominate day to day life. With inadequate health infrastructure and unaffordable quality healthcare, numerous alternative, community based models designed to build health capacity have been developed and implemented in recent decades [1-4].

Community health workers (CHWs) are local health workers who do not have a professional degree or license and who serve as health representatives within their communities. CHWs currently fill an extremely large gap in LMICs by providing health services and resources, including life-saving triage and treatment modalities to help connect sick patients from community to clinic [5, 6, 8]. Several studies have cited how CHWs are effective in meeting a diverse array of essential community health needs [1, 6]. However, with limited resources and training, significant challenges to providing structure and sustainable methodology for CHW program development exist. This, coupled with limited data on effective pedagogy for training CHWs in remote regions, has created a dire need for standardization to establish effective, sustainable, and community-rooted public health initiatives utilizing CHWs [8-10].

Program models employing methodologies that transition learner to instructor over a defined period of time have successfully demonstrated that this type of train-the-trainer methodology not only empowers those transitioning to instruct but ensures continued competency in learning objectives [2-4]. With this transitional model, sustainability is enhanced as the resource input is diminished, while local community leaders establish the infrastructure needed to develop local health capacity, including training and treatment sites [2-4].

Several new CHW training models have integrated the use of mobile health (mHealth) technology. mHealth, a subset of electronic health (eHealth), consists of digital health information applied to mobile health technology [11-13]. With the widespread use of cellular phones, and more recently, tablet computers, information exchange across some of the most desolate, limited resource settings in the world is now feasible [14-15]. With improved accessibility, the use of health information on mHealth can augment CHW training programs with improved training, monitoring, and treatment protocols. Further, with limited available resources in many rural programs, the application of mHealth can provide recurrent training and augment retention with repeated lessons in a setting without in-person instructors. *Few studies to date have evaluated the effectiveness of combining train-the-trainer and mobile health methodologies for community-based health training programs in LMICs*. This study aimed to address the effectiveness of combining a train-the-trainer model with mHealth by assessing medical skill competency over time and comparing initial trainee scores from two sets of instructors: trained CHWs and certified health professionals.

## Materials and Methods

### Laro Kwo Project

The Laro Kwo Project (LKP) was founded in 2016 to establish a self-sustaining, participatory CHW training program led by community members in Northern Uganda. Laro Kwo, an Acholi name agreed upon by local villages in Northern Uganda, translates to ‘saving lives’ in English. The development of the LKP was facilitated by the nonprofit organizations Northern Uganda Medical Mission (NUMEM) and MGY (the name MGY represents the formula for potential energy in physics). NUMEM operates a health clinic in Pader, Northern Uganda, and MGY is a multidisciplinary team primarily composed of physicians and engineers based in the United States who provide resources for mobile health program development in limited resource settings.

The LKP was initiated after one year of discussions among village leaders, local elected officials, the Uganda Ministry of Health, and both NUMEM and MGY. Village leaders recommended a total of 25 CHWs among their communities to begin training under the LKP. Preliminary meetings with the above stakeholders and community members identified specific health concerns that formed a framework for the training program curriculum. Once finalized, MGY drafted educational materials, including videos and quizzes of the requested training materials and presented them to stakeholders. Once approved by all stakeholders, NUMEM facilitated translations of materials and set up a date and time to begin the first training session with in-person didactics. The training curriculum focused on emergency triage, prehospital emergency care, and public health.

### Medical Skill Retention

In year 1 of the LKP, the 25 selected CHWs completed a five-day course which was held at the health ministry quarters in Kilak sub-county in Northern Uganda. Present for the training was a team of MGY physicians and engineers, NUMEM physicians and coordinators, ministry of health officials, elected local representatives and local village leaders. After completing didactic training on specified skills (table 1), each CHW completed a 1-on-1 skill competency exam the last day of training. The same instructor evaluated one skill for consistency. Skill competency exams had a maximum score of 42 points, or 100%.

**Table 1.**
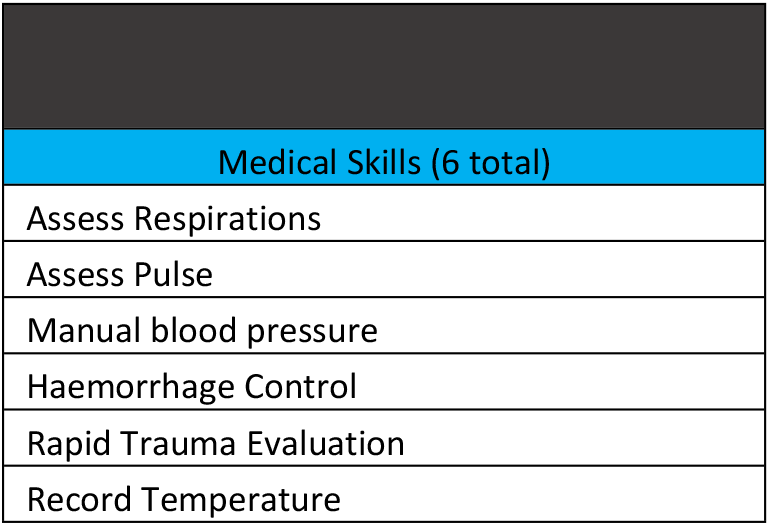
Annual Medical Skills Completed by CHWs.

| Table 1. Annual Medical Skills Completed by CHWs |
| --- |
| Medical Skills (6 total) |
| Assess Respirations |
| Assess Pulse |
| Manual blood pressure |
| Haemorrhage Control |
| Rapid Trauma Evaluation |
| Record Temperature |

Annual trainings were repeated the following two years with reassessment of skills after this initial training. During each annual training, a new cohort of 25 CHWs was added. After two years of passing >80% on skill competency, CHWs were offered promotion to advanced CHW and instructor status. After the second annual training session, a cohort of six CHWs chose to transition to instructors and revised the core training materials on the mHealth application. This included video demonstrations for each skill, which they proceeded to instruct at the next (year 2) annual training.

A three-year prospective observational study was completed to evaluate changes in competency exam scores for specific medical skills. We hypothesized that with personal tablet computers loaded with a LKP tailored mobile health application, the original 25 CHWs would maintain annual medical skill competency scores without significant lapse in retention. Further, we hypothesized that by using a train-the-trainer model with mHealth, advanced CHWs could transition to instructor status and disseminate material equally effectively.

### Statistical Analysis

CHWs’ skill competency exam scores are presented as mean (95% CI). The original 25 CHW scores were modeled over time using mixed linear regression. The original CHW initial scores were compared with the initial scores from new CHWs who started in year 3 using Student’s t-test.

### Ethics Statement

Ethics approval for the study design, recruitment, and methods was obtained from A.T. Still University School of Osteopathic Medicine Institutional Review Board and the Uganda National Council for Science and Technology Ethics Committee. Written consent was obtained from each participant in study. During CHW recruitment, it was clarified that there was no risk in declining to participate in study.

## Results

Modeled mean skill competency exam scores for the original CHWs were 80.3% (76.7%, 84.0%) for year 1, 90.6% (87.5%, 93.6%) for year 2, and 88.7% (86.8%, 90.7%) for year 3 (Table 2). There was evidence for a meaningful difference in scores by year in the overall comparison (p <0.0001) as well as for pairwise comparisons (Table 2), with an increase between year 1 and year 2 (p <0.0001) and between year 1 and year 3 (p <0.0001). There was no evidence for a meaningful difference in scores between years 2 and 3 (p=0.26).

**Table 2.** Original CHW Scores.

| Table 2. Original CHW Scores |  |  |  |  |  |
| --- | --- | --- | --- | --- | --- |
| Effect | year | Estimate | Lower | Upper | P Value |
| year | 1 | 0.8033 | 0.7669 | 0.8398 | - |
| year | 2 | 0.9056 | 0.8751 | 0.9360 | <.0001 |
| year | 3 | 0.8872 | 0.8678 | 0.9067 | <.0001 |

Average CHW competency scores for first training with years 2 and 3 representing trainings by advanced CHWs. P-values represent difference between first year scores.

Comparing the original CHW initial scores with initial scores from CHWs who started in year 3 (“new CHWs”), the new CHWs had a higher mean initial score (Table 3) of 90.7% (89.0%, 92.4%) than the original CHW mean score of 80.3% (76.7%, 84.0%; p<0.0001). The scores from year 3 represent scores from CHWs that were trained by advanced CHWs, completing the train-the-trainer cycle.

**Table 3:** A comparison between original CHWs scores (year one) and new CHWs trained by advanced CHWs (year three)

|  | CHW group |  |  |  |
| --- | --- | --- | --- | --- |
|  | *new*<br>(N = 25) | *original*<br>(N = 25) | *Total*<br>(N = 50) | *P Value* |
| Total score |  |  |  |  |
| Mean (SD) | 38.10 (1.70) | 33.74 (3.83) | 35.92 (3.67) | <0.001*** |
| Median (IQR) | 38.0 (36.5 – 39.5) | 34.0 (31.0 – 37.0) | 36.8 (34.0 – 39.0) | <0.001*** |
| Percentage |  |  |  |  |
| Mean (SD) | 0.91 (0.04) | 0.80 (0.09) | 0.86 (0.09) | <0.001*** |
| Median (IQR) | 0.9 (0.9 – 0.9) | 0.8 (0.7 – 0.9) | 0.9 (0.8 – 0.9) | <0.001*** |

## Discussion

This study demonstrates that the use of train-the-trainer with mHealth is an effective model for sustainable community program development in LMICs. After reconstructing core medical education materials and transitioning over three years to instructor, trained CHWs effectively delivered the training to 25 new CHWs. The scores of new CHWs averaged higher than previous trainings implemented by local and foreign health professionals, with sustained high competency scores in years 2 and 3 as advanced CHWs assumed leadership of the program.

CHWs provide a critical resource to community members that can influence patient outcomes in LMICs. Particularly in regions such as Northern Uganda, where no regional hospital exists, careful triage of community members who may require hospitalization can be life-saving [16]. Critical to this role is the maintenance of basic skill competency for effective triage evaluation. Many studies recognize the effectiveness of CHWs in health programs but fail to recognize the need for sustainability and note limited evidence for best practices in training modality [7-8]. A recent systematic review of CHW training programs focusing on LMICs highlights the scarcity of evidence for CHW training impact and the need for investigation of effective training modalities [8]. Further, this review emphasized the importance of community embeddedness to program success, and how increased motivation and confidence can lead to improved program services. The results from this study support the tenet that community focused training programs improve confidence and motivation, which link to program success.

Similarly, recent reviews of mHealth and interventions among CHWs outline the effectiveness of training CHWs with mHealth. However, these studies note several challenges, such as lack of culturally relevant mHealth interventions, lack of consistent methodology to assess outcomes, and need for further studies evaluating effective training modalities [8, 17-18]. This study addresses some of these concerns and highlights how a community-based training program can sustain effective health training in limited resource settings. Moreover, and fundamental to this concept, the study demonstrates that combining train-the-trainer with mHealth ensures cultural relevance to education materials while driving motivation and self-empowerment. Future studies replicating this methodology will explore combinations of training modalities that are community-based and culturally tailored to increase sustainability. Additional studies addressing clinical outcomes of various CHW program models are necessary to validate a positive health impact of programs such as the LKP.

## Data Availability

Data collected, VHT scores, are deidenfitied and deposited publicly on Open Science Framework. https://osf.io/mhd94/?view_only=6fffe121df6a4172a12cc0d17f0f2b64 Ebbs, D. (2022, October 22). The Laro Kwo Project: A train the trainer model combined with mobile health technology for Community Health Workers in Northern Uganda. Retrieved from osf.io/mhd94

## Acknowledgments

The authors would like to acknowledge the time and commitment to training organized and implemented by the nonprofit organizations MGY and Northern Uganda Medical Mission; and the Uganda Ministry of Health. Furthermore, we would like to acknowledge David Tseng and Sam Waggoner for the development of a community tailored mobile health application. Finally, we would like to acknowledge the hard work and commitment of the 75 community health workers who continuously serve their communities as critical health resources.

## References

1. Singh P, Sachs JD. 1 million community health workers in sub-Saharan Africa by 2015. The Lancet. 2013 Jul 27;382(9889):363–5.

2. Rodriguez NM, Casanova F, Pages G, Claure L, Pedreira M, Touchton M, et al. Community-based participatory design of a community health worker breast cancer training intervention for South Florida Latinx farmworkers. PloS one. 2020 Oct 19;15(10):e0240827.

3. Vernon MM, Albashir G, Sojourner SJ, Moore JX, Looney SW, Tingen MS. Using a “train-the-trainer” approach with urban and rural minority community health workers to implement the cancer-Community Awareness Access Research and Education (c-CARE) Project. Cancer Research. 2021 Jul 1;81(13_Supplement):2553-.

4. Yu X, Pendse A, Slifko S, Inman AG, Kong P, Knettel BA. Healthy people, healthy community: evaluation of a train-the-trainers programme for community health workers on water, sanitation and hygiene in rural Haiti. Health Education Journal. 2019 Dec;78(8):931–45.

5. Kane SS, Gerretsen B, Scherpbier R, Dal Poz M, Dieleman M. A realist synthesis of randomised control trials involving use of community health workers for delivering child health interventions in low and middle income countries. BMC health services research. 2010 Dec;10(1):1–7.

6. Thapa R, Thrift A, Mishra SR, Zaman SB, Zengin A. 1433 Community Health Workers for non-communicable diseases: a systematic review and meta-analysis. International Journal of Epidemiology. 2021 Sep;50(Supplement_1):dyab168–659.

7. Scott K, Beckham SW, Gross M, Pariyo G, Rao KD, Cometto G, Perry HB. What do we know about community-based health worker programs? A systematic review of existing reviews on community health workers. Human resources for health. 2018 Dec;16(1):1–7.

8. Kok MC, Dieleman M, Taegtmeyer M, Broerse JE, Kane SS, Ormel H, et al. Which intervention design factors influence performance of community health workers in low-and middle-income countries? A systematic review. Health policy and planning. 2015 Nov 1;30(9):1207–27.

9. Israel BA, Schulz AJ, Parker EA, Becker AB, Allen AJ, Guzman JR, et al. Critical issues in developing and following CBPR principles. Community-based participatory research for health: Advancing social and health equity. 2017 Oct 5;3:32–5.

10. Omer K, Mhatre S, Ansari N, Laucirica J, Andersson N. Evidence-based training of frontline health workers for door-to-door health promotion: a pilot randomized controlled cluster trial with Lady Health Workers in Sindh Province, Pakistan. Patient education and counseling. 2008 Aug 1;72(2):178–85.

11. Early J, Gonzalez C, Gordon-Dseagu V, Robles-Calderon L. Use of mobile health (mHealth) technologies and interventions among community health workers globally: a scoping review. Health promotion practice. 2019 Nov;20(6):805–17.

12. Mahmood H, Mckinstry B, Luz S, Fairhurst K, Nasim S, Hazir T, Respire Collaboration. Community health worker-based mobile health (mHealth) approaches for improving management and caregiver knowledge of common childhood infections: A systematic review. Journal of global health. 2020 Dec;10(2).

13. Ebbs D, Hirschbaum JH, Mika A, Matsushita SC, Lewis JH. Expanding Medical Education for Local Health Promoters Among Remote Communities of the Peruvian Amazon: An Exploratory Study of an Innovative Program Model. Advances in Medical Education and Practice. 2020;11:215.

14. Betjeman TJ, Soghoian SE, Foran MP. mHealth in sub-Saharan Africa. International journal of telemedicine and applications. 2013 Oct;2013.

15. Colaci D, Chaudhri S, Vasan A. mHealth interventions in low-income countries to address maternal health: a systematic review. Annals of global health. 2016 Sep 1;82(5):922–35.

16. Udhs I. Uganda demographic and health survey. Uganda Bureau of Statistics, Kampala Uganda. 2016.

17. Abreu FD, Bissaco MA, Silva AP, Boschi SR, Scardovelli TA, Santos MF, et al. The use and impact of mHealth by community health workers in developing and least developed countries: a systematic review. Research on Biomedical Engineering. 2021 Sep;37(3):563–82.

18. Schoeman F. Digital tools for training frontline health workers in low and middle-income countries: A systematic review (Master’s thesis, Faculty of Health Sciences).

